# Study protocol: A national cross-sectional study on psychology and behavior investigation of Chinese residents in 2023, PBICR

**DOI:** 10.1101/2024.03.28.24305038

**Authors:** Diyue Liu, Siyuan Fan, Xincheng Huang, Wenjing Gu, Yifan Yin, Ziyi Zhang, Baotong Ma, Ruitong Xia, Yuanwei Lu, Jingwen Liu, Hanjia Xin, Yumeng Cao, Saier Yang, Runqing Li, Han Li, Ji Zhao, Jin Zhang, Zheng Gao, Yaxin Zeng, Yixiao Ding, Zhuolun Ren, Yan Guan, Na Zhang, Jia Li, Yan Ma, Pei Wei, Jingjing Dong, Yajing Zhou, Yong Dong, Yan Qian, Chen chen, Yujie Zhao, Yimiao Li, Yujia Zheng, Rongyi Chen, Xiaomeng Li, Yuke Han, Yaoyao Xia, Huixin Xu, Zhaolin Wu, Mingyou Wu, Xinrui Wu, Junyi Hou, Yuelai Cai, Xiaofan Dai, Wenbo Li, Ting Nie, Chongzhe Zhang, Xiaoya Wang, Dan Li, Siyao Yan, Chenxi Liu, Xinyue Zhang, Lei Shi, Haomiao Li, Feng Jiang, Xiaoming Zhou, Xinying Sun, Yibo Wu the Psychology and Behavior Investigation of Chinese Residents project team

## Abstract

**Background:** This study protocol specifies the primary research line and theoretical framework of the 2023 Survey of the Psychology and Behavior on Chinese Population. It aims to establish a database focused on psychology and behavior among Chinese residents through a multicenter, large-sample cross-sectional survey to provide strong data support for research and development in related fields. It will track the public’s physical and psychological health more comprehensively and systematically.

**Methods:** The study was conducted from 20 June 2023 to 31 August 2023, using stratified and quota sampling methods, and a total of 150 cities, 202 districts and counties, 390 townships/streets, and 800 communities/villages were surveyed (excluding Taiwan, which were selected from 23 provinces, five autonomous regions, and four municipalities directly under the central government in China). The questionnaires were distributed face-to-face by trained surveyors. The questionnaires included basic information about the individual, personal health status, basic information about the family, the social environment in which the individual lives, psychological condition scales, behavioral level scales, other scales, and attitudes towards topical social issues. Supervisors conducted quality control during the distribution process and returned questionnaires, logically checked and cleaned for data analysis.

**Discussion:** Data collection has been finished, and scientific outputs based on this data will support the development of health promotion strategies in China and globally. In the aftermath of the pandemic, it will guide policymakers and healthcare organizations to improve their existing policies and services to maximize the physical and mental health of the Chinese population.

## 1 Background

Mental health is an integral and essential component of health. Mental health is a state of mental well-being that enables people to cope with the stress of life, realize their abilities, learn well and work well, and contribute to their community(*Mental Health*, 2022). An important implication of this definition is that mental health is more than just the absence of mental disorders or disabilities(*Health and Well-Being*, n.d.). Health behaviors include health-enhancing behaviors and health-impairing behaviors. According to the World Mental Health Report 2022: The Transition to Access to Mental Health Services for All, about 9.7 million people worldwide suffer from mental disorders. It is estimated that the incidence of anxiety disorders and depression increased by more than 25% worldwide during the first year of the Corona Virus Disease 2019 (COVID-19) pandemic(Najafipour et al., 2021; *World Mental Health Report*, 2022). Reports significant and consistent but modest associations between COVID-19 infection and increased rates of psychiatric disorders(Weich, 2022). The risk of depression was detected in 10.6%, and the risk of anxiety was detected in 15.8% of Chinese nationals(Shen, 2023). According to the first national survey on psychological distress during the COVID-19 epidemic in China in 2020, 35% of respondents experienced distress, including anxiety and depression. According to changing trends in the prevalence of mental disorders, we can understand that the COVID-19 pandemic is a significant stressor for the public. Health behaviors were defined in 2011 by B. Renner as “Health behaviors belong to the broader category of health-directed activities. They are aimed at preventing or detecting illnesses at an asymptomatic stage.” (*World Mental Health Report*, 2022),Health behaviors such as diet quality, physical activity, alcohol use, smoking, sleep and sitting-time behaviors, in particular, sleep, have a strong association with mental health.(Oftedal et al., 2019). The survey conducted this year also covered some special dimensions of access, such as children’s adverse experiences (Felitti et al., 1998). Since the COVID-19 pandemic, tobacco smoking cessation and quit attempts have increased, the prevalence of sleep disorders has increased, and the epidemic has profoundly impacted health behaviors and mental health(McBride et al., 2021; The Lancet, 2022).

Mental health is determined by the complex interplay of individual, family, community, and structural factors (sociocultural, geopolitical, and environmental factors), especially health behavior factors. These factors vary with time and space, and everyone’s experiences are not the same(*World Mental Health Report*, 2022; Wright et al., 2022). Health behaviors and mental health often interact with each other. Physical activity affects the generation of psychological problems(Mukhtar, 2020; Shaw et al., 2023). Regular physical activity is associated with better mental health(Stubbs et al., 2018). Besides, Social factors, including social activities, influence the generation of psychological problems. Exposure to unfavorable social, economic, geopolitical and environmental circumstances-including poverty, violence, inequality(d’Elia et al., 2022) and environmental deprivation-also increases people’s risk of experiencing mental health conditions. Mental health can also affect behavior accordingly(Kunzler et al., 2020). Because of the interaction between psychological health and health behaviors, and the impact of psychological health and health behaviors on physical health(Aoki et al., 2020), many countries have proposed a series of measures to promote the population’s psychological health and health behaviors. For example, the UK government has proposed a psychological health promotion strategy that covers the entire population, and the Japanese government has shared the extent of its past mental health promotion efforts and future priorities on the government website(*Japan Health Policy NOW – Mental Health*, 2021; *The Mental Health Strategy for England*, 2011). These national mental health promotion strategies are based on nationally representative data, so it is essential to establish a high-quality database of mental health and health behavior surveys.

In Chinese studies, most surveys focused only on common factors, such as anxiety and depression, social factors, and lifestyle, and the study population was also relatively single. According to the first national survey on psychological distress in the COVID-19 epidemic in China starting in 2020, 35% of respondents experienced distress, including anxiety and depression(The Lancet, 2022). Cao L(Lu et al., 2022) et al., Yu LL(Yu et al., 2022) et al., Wu Y(Wu & Peng, 2020) et al., and Liu C(Cui et al., 2015) et al. conducted research studies on depression, anxiety, and their associated factors for older adults in Shenzhen, medical and non-medical staff during the new coronavirus pneumonia epidemic, hospitalized patients with severe novel coronavirus pneumonia, and hypertensive co-morbidities in Beijing and Jilin provinces, respectively, and Wei CL (Wei et al., 2023)et al., He YY(He et al., 2021) et al., Yang H(Yang et al., 2020) et al., and Sun J(Sun & Lyu, 2020) et al. studied social factors, such as residents’ sense of social justice, income inequality and social capital, social participation, about physical and mental health, Liang XY(Liang et al., 2020) et al., Chen Y(Chen et al., 2022) et al., Zhao J(Jin et al., 2021) et al. studied the relationship between behavior and mental health, such as short video use among female college students, unhealthy lifestyles of Chinese psychiatrists, risky sexual behaviors among Chinese college students, respectively. Cross-sectional studies of mental health and health behaviors in China have investigated a relatively homogeneous set of variables, often ignoring the influence of an individual’s literacy, individual family factors, and individual past experiences on existing mental health status and established health behaviors. This makes it difficult for studies based on these surveys to discuss individual psychological problems and the causes of poor health behaviors more fully. Moreover, previous studies have often focused on specific groups when focusing on mental health and health behaviors, making it difficult to provide a scientific basis for population-wide health promotion.

In summary, there is a lack of recent research on the current situation of many aspects of public mental and behavioral health in China, covering multiple groups, to provide academics and policymakers with relevant data and references for empirical research. Psychology and Behavior Investigation of Chinese Residents (PBICR) has surveyed for three consecutive years(Y. Wang et al., 2022; *2021 China Family Health Index Survey General Report*, 2023; *Chinese People’s Health Status Report 2020*, 2021), with the variables surveyed retained and replaced in each year, this year was a survey conducted after the COVID-19 pandemic, aiming to establish an extensive multicenter, repetitive, nationwide cross-sectional database of mental and behavioral health, which enable researchers to probe the correlation between different psychological and behavioral variables and different populations, and explore the public’s psychological and behavioral trends by applying data accumulated over three years,, enrich and deepen theoretical and practical research in the field of mental health, helping to promote the strategy of “Healthy China”(W. Wang & Zakus, 2016) on all fronts, and respond to the World Health Organization(WHO) Comprehensive Action Plan on Mental Health 2013-2030.

## 2 Methods

### 2.1 The study design and setting

This cross-sectional survey was collaboratively launched by the School of Public Health at Peking University, the Institute of Healthy Yangtze River Delta at Shanghai Jiao Tong University, and Shandong Provincial Hospital. It was conducted from June 20, 2023, to August 31, 2023, encompassing 150 cities, 202 districts and counties, 390 township streets, and 800 community villages across 23 provinces, five autonomous regions, and four municipalities directly under the Central Government of China. Data was also collected in Hong Kong and Macau regions (excluding Taiwan). The study received approval from the Biomedical Ethics Committee of Shandong Provincial Hospital (Grant No.: SWYX: NO.2023-198) on May 5, 2023. Subsequently, it was filed in the National Health Security Information Platform (Record No.: MR-37-23-017876) and officially registered in the China Clinical Trials Registry (Registration No.: ChiCTR2300072573).

### 2.2 Study participants and data collection

This study is a cross-sectional study with an unlimited number of subjects, and the final minimum sample size required for this study was determined according to this sample size formula.

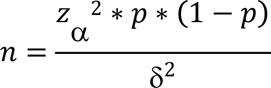

Where the prevalence refers to the results of the meta-analysis by Krishnamoorthy(Krishnamoorthy et al., 2020) et al., the prevalence of depression in the general population of China is 24%. The given formula establishes the probability of Type I error (α) as 0.05, while the acceptable error is determined to be 0.02. Consequently, the minimum sample size is computed as 1,752. Given that the planned sample size for this study surpasses the minimum requirement, the allocation of sample size for each province is based on the proportion of its respective population.

The study was conducted nationwide, considering the population proportions provided by the 7th National Census, encompassing 23 provinces, five autonomous regions, four municipalities directly under the Central Government, as well as Hong Kong and Macau. Sampling proportions were determined accordingly. From each province/autonomous region/municipality directly under the central government, a minimum of 500/1,000/1,500/2,000/2,500/3,000 individuals were sampled, except Hong Kong and Macau, where 200 individuals were sampled each. The total estimated sample size for the study was 40,000 individuals. The sampling process included selection at various administrative levels, such as city, district, county, township, street, and community/village levels. Additionally, sampling was carried out at the individual level, considering both gender and age as quota attributes. The proportion of the sample size expected to be recovered in each province is shown in **Figure 1**.

**Figure 1.**
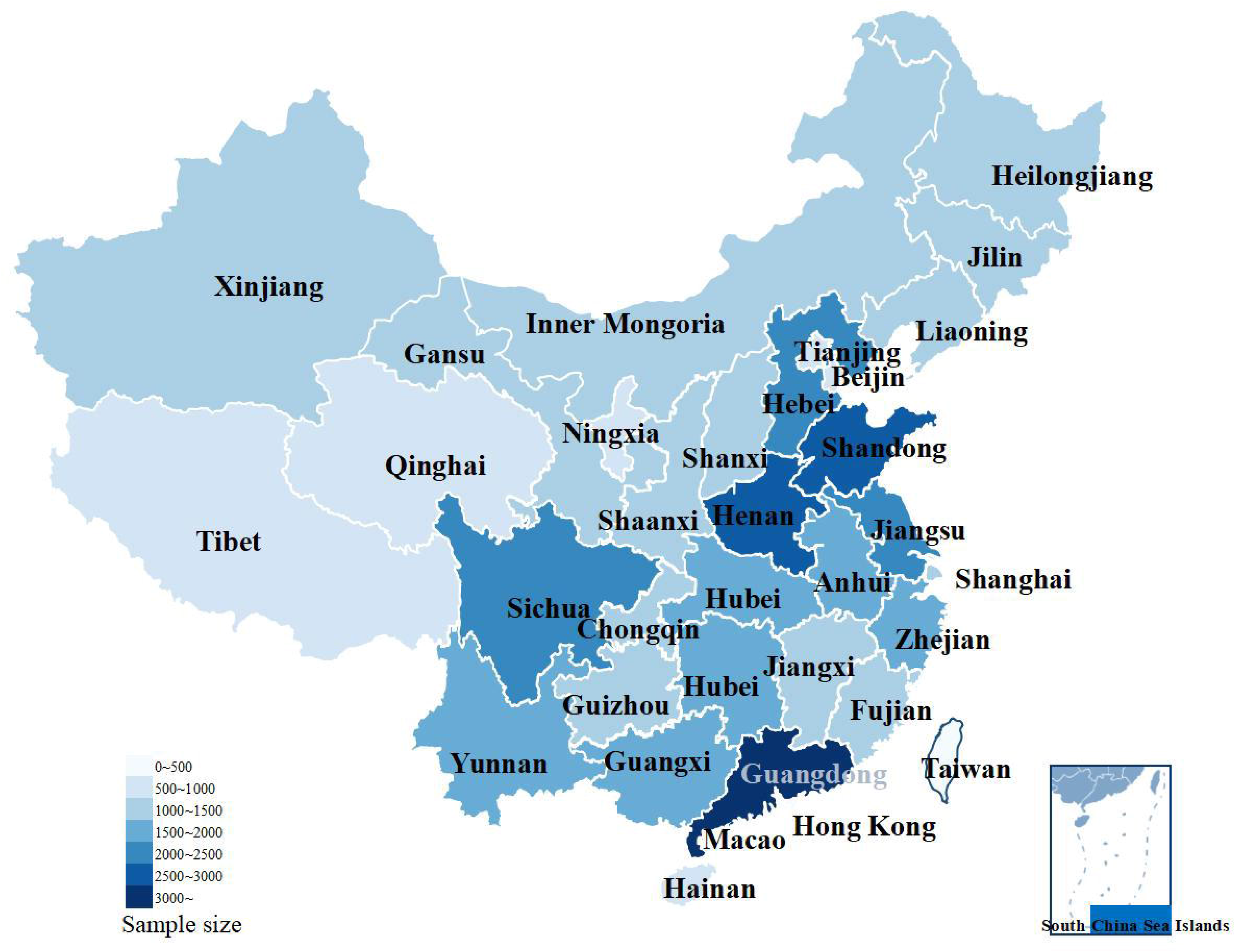
Sampling weight by province.

A multi-stage sampling approach was adopted for this study. During the initial phase, the research team directly encompassed 23 provinces’ capital cities, five autonomous regions, and four municipalities (Beijing, Tianjin, Shanghai, and Chongqing). Additionally, 1-12 cities were selected from each non-capital prefecture within every province, municipality, and autonomous region, resulting in 150 cities. The number of cities selected in each province is determined by the weight of the population size in that province, with each province including its capital city and the rest of the cities selected by convenience sampling. Moving on to the second stage, 6-36 rural and 312 urban communities were randomly selected from each of the 150 cities above, yielding a cumulative total of 800 communities. In the third stage, a quota sample of 150 urban residents was drawn, considering the findings of the “7th National Census in 2021.” This sample specifically accounted for attributes such as gender and age. For instance, if a province required 1,000 data points, a balanced ratio of men to women at 1:1 would be upheld, entailing 500 men and no fewer than 500 women. Moreover, the urban areas would comprise at least 400 individuals, while the rural areas would include at least 600 individuals.

Due to time constraints, the project team was not able to shortlist a suitable candidate to conduct the survey in Taiwan. At the same time, due to financial limitations, the project team was not able to send a qualified person in charge to Taiwan to complete the survey.

The survey included Chinese permanent residents who hold citizenship of the People’s Republic of China and have an annual departure time of ≤ 1 month. Eligible participants had to be at least 18 years old (born before August,31, 2005), possess the ability to comprehend each item in the questionnaire and be capable of independently completing the questionnaire or doing so with the assistance of an investigator. All participants volunteered to participate in the study and were required to provide informed consent by signing a consent form. Exclusions encompassed individuals with confusion, psychiatric abnormalities, or cognitive impairment. Moreover, those who had previously participated in similar studies or expressed a lack of interest in participating in this particular study were also excluded.

### 2.3 Quality control

The research team implemented quality control measures across six stages in this survey. These stages encompassed questionnaire design, pre-survey activities, investigator training, questionnaire distribution, logic checking, and data processing. During the questionnaire design stage, the scale design team carefully assessed the logical structure of the questionnaire and diligently identified any evident errors in its design. In the pre-survey stage, the quality control team conducted three pre-surveys to identify any issues related to the questionnaire setup. Feedback on these issues was provided and promptly communicated to the scale design group for timely revisions. In the investigator training stage, the training and coordination group ensured the selection of high-quality provincial leaders and investigators or investigator teams. Through three comprehensive survey trainings, the team enhanced the investigators’ understanding of the project and their research skills, guaranteeing that their work adhered to the rigorous standards established by the research team. In the questionnaire distribution stage, provincial leaders and investigators or investigator teams were required to adhere to the survey standards provided by the research team when distributing the questionnaires. Please comply with these standards to avoid exclusion from the pool of qualified questionnaires. During the logic-checking phase, the research team screened the collected questionnaires once a week and eliminated those that needed to meet the logic-checking rules set in advance. Lastly, during the data processing stage, upon completion of the survey, the research team diligently checked and cleaned the questionnaire data by stringent quality screening standards.

In order to ensure the quality control of every aspect of the project, we have set up a complete organizational framework to execute every aspect of the project. The organizational framework of this survey project follows a general manager system, wherein each team is led by an individual who oversees and coordinates their collaborative efforts to complete the survey project. The research team comprises five distinct research groups, namely the expert committee, survey group, training and coordination group, scale design group, and quality control group:

1. The expert committee comprises invited experts in psychology, behavior, public health, and statistics. Their role is to provide expertise and guidance throughout the project.
2. The survey group comprises provincial heads, investigator teams, and investigators. Each provincial administrative region is assigned a research team member to supervise operations. The provincial heads assume responsibility for recruiting, training, organizing, and coordinating of investigators or investigator teams within their respective provinces.
3. The training and coordination group focuses on investigator training, ethical review submission, study protocol registration, and survey protocol writing.
4. The scale design group is primarily responsible for screening questionnaire variables, developing original scales, incorporating international scales, simplifying scales, and establishing logic settings for skip questions.
5. The quality control group is pivotal in expert consultations, pre-survey activities, ensuring sampling quality, and verifying questionnaire logic. The supervisor responsible for questionnaire distribution and retrieval followed the study throughout.

#### 2.3.1 Questionnaire design phase

The questionnaire design phase commenced with the research team conducting in a comprehensive scientific review of relevant literature and scholarly works. To ensure the questionnaire’s efficacy, the research team organized online expert consultation and discussion from March to June 2023. Forty-two experts, possessing esteemed credentials in fields such as social medicine, behavioral epidemiology, psychology, health education, health statistics, health service management, humanistic medicine, journalism and communication, clinical medicine, pharmacy, nursing, sociology, and philosophy, were invited to evaluate the quality of the initial draft questionnaire and provide recommendations for revisions. Subsequently, the research team meticulously incorporated the experts’ feedback, resulting in a refined version of the questionnaire. To validate its effectiveness, the research team conducted three rounds of pre-survey research based on the revised questionnaire.

At the early stage of questionnaire design, the relevant workers of the project group retrieved a large number of English literature libraries while referring to the questioning methods of well-known databases at both domestic and foreign countries to select a large number of scales, and all the scales included in the survey were determined through multiple rounds of expert consultation and voting by forty-two experts.

#### 2.3.2 Pre-investigation phase

The pre-investigation phase of the survey consisted of three rounds conducted during three distinct periods: June 5 to June 8, June 10 to June 13, and June 15 to June 18, 2023. The sampling method employed in the pre-survey stage was fixed with fixed attributes, consistent with the method employed in the official survey. The pre-survey sample sizes for the three rounds were 100, 100, and 200, respectively. The research team collected and compiled respondents’ feedback and modifications throughout the pre-survey period. They extensively discussed and revised the questionnaire based on the results of statistical analyses about questionnaire reliability. The revised questionnaire underwent subsequent review by the expert committee and was finalized. It is important to note that the questionnaires collected during the pre-survey phase were not included in the final study analysis.

#### 2.3.3 Investigators’ training process

The research team publicly released the “Announcement on the Recruitment of Provincial Leaders for the National Cross-Section Survey” online and conducted an initial screening of the received resumes. The primary screening criterion for provincial leaders was relevant survey experience and scientific research background. Subsequently, the research team notified the applicants for an interview, which assessed their scientific research proficiency, managerial competence, communication skills, decision-making abilities, stress tolerance, and adaptability to language environments. Applicants who pass the interview will be formally appointed as provincial leaders, with at least two leaders assigned per province, municipality, or autonomous region. The research team conducted intensive training sessions lasting 1-2 hours on May 13, May 14, and May 28, 2023, to ensure that the provincial leaders possess comprehensive knowledge and a deep understanding of this survey project. Once the training of provincial leaders is completed, the research team will assist them in recruiting investigators or investigation teams in each province. The provincial leaders will be responsible for organizing investigators’ recruitment, screening, and training. Those investigators who pass the training assessment will be assigned specific investigation tasks within their designated areas.

#### 2.3.4 Questionnaire Distribution Process

The research team adhered to scientific design principles and statistical methodology requirements to ensure rigorous control over the questionnaire distribution process, minimizing potential biases in data collection. The project team asked all investigators to distribute the return questionnaires face-to-face. The data collection flow can be visualized in **Figure 2** presented below. Initially, the team-mandated investigators applied a coding system to each questionnaire. The coding principle aimed to facilitate traceability of questionnaires to the corresponding subjects, investigators or investigation teams, and provincial leaders. Questionnaires failing to adhere to the coding rules were deemed invalid. Additionally, before commencing the official survey, provincial leaders reiterated potential challenges to investigators multiple times, aiming to maximize the validity of each collected questionnaire. Subsequently, once the official survey commenced, the research team established weekly communication sessions with provincial leaders via Tencent Meeting, every Sunday at 8:00 p.m.(from 20 June 2023 to 31 August 2023). These sessions encompassed providing feedback on the quality of the week’s questionnaires, addressing existing issues, and responding to inquiries from provincial leaders. In case of any problems encountered during the investigation process, provincial leaders were expected to rectify them according to the research team’s guidelines. Real-time discussions were required to propose and deliberate upon potential solutions in the event of disagreements.

**Figure 2.**
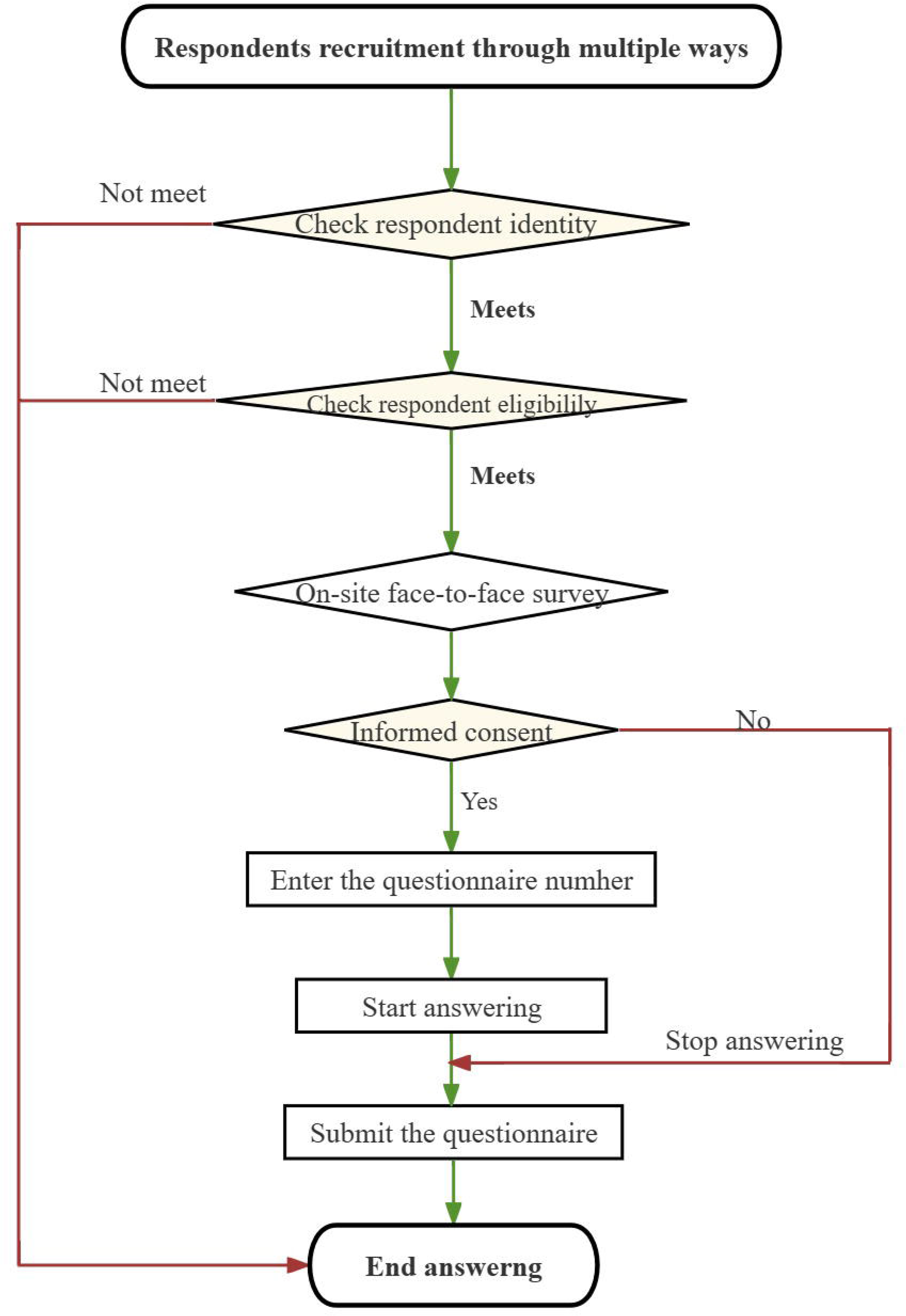
Data collection flow chart.

#### 2.3.5 Questionnaire retrieval and analysis process

The research team will establish stringent criteria for screening valid questionnaires before the official survey commencement. Each criterion will undergo logical verification by two investigators consecutively to ensure the accuracy and validity of the assessment. Questionnaires must meet the criteria to be deemed valid. The questionnaire screening criteria include: ① Questionnaires with response times below 600 seconds. ② Questionnaires that do not pass the logic check.(such as selecting “have religious beliefs” in question 5 or selecting “member of the Communist Party of China,” “reserve member of the Communist Party of China,” or “member of the Communist Youth League” in question 9; Questionnaires where the selected “highest level of education” in question 11 is higher than the selected “current study period” in question 12; Questionnaires where the “current place of residence” in question 17 matches the “place of birth” in question 21, et.al③Incompletely filled out questionnaire(Questionnaires with more than 20% missing values).④Questionnaires that are filled out repeatedly. ⑤Questionnaires with the same selected options or regular patterns(Straight-lining participants will be judged to have completed all the scales with the same answers.). Additionally, the research team will conduct a statistical analysis of the questionnaires. If any irregular values are identified, the team will trace them back to the original questionnaires or consult with the investigators to verify the information. The study flow can be visualized in **Figure 3** presented below.

**Figure 3.**
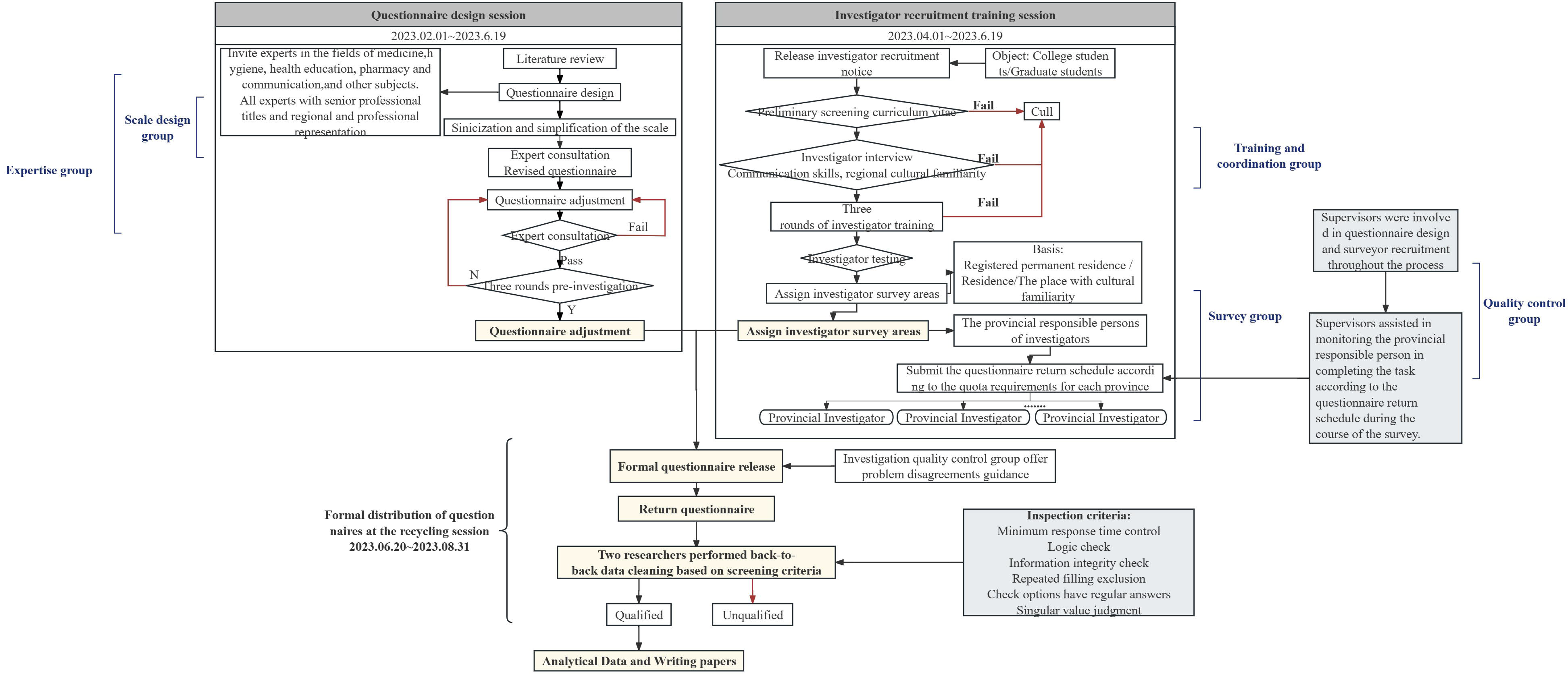
Study flow chart.

### 2.4 Measurements

In addition to informed consent and questionnaire numbering, the questionnaire for this study consisted of seven main areas: fundamental personal details, personal health condition, family demographic information, psychological level scales, behavioral level scales, other scales(They are made up of maturation scales, self-administered scales and questionnaires), and attitudes toward socially topical issues. Detailed information of the scales is shown in **table 1**.

#### 2.4.1 Fundamental personal details

This section encompasses 23 inquiries regarding various aspects, including gender, ethnicity, religion, height, weight, waistline, political status, occupation and occupational status, the highest level of education attained, academic background (in terms of schooling, college, and major), migration details (birthplace, current residence, length of residency, and place and time of migration), and preferred hand.

#### 2.4.2 Personal health condition

This section comprises ten questions about chronic disease diagnoses, classification of hypertension, complications associated with hypertension, types of diabetes mellitus, respiratory disease classification, urinary disease classification, digestive disease classification, tumor types, status of injury events, and experiences of life events.

#### 2.4.3 Family demographic information

This section includes 21 questions addressing permanent residence, household registration, marital status, family type, romantic relationship status, living arrangements (whether living alone, with a spouse, with spouse’s parents, or with parents), number of children, living situation with children, age of youngest and oldest child, number of siblings, living situation with siblings, residential area, number of residential properties, debt status, monthly per capita household income, and methods of medical expense coverage.

## 3 Discussion

The primary objective of this study is to offer an all-encompassing and methodical comprehension of the Chinese populace’s present mental health and health behaviors. This will be achieved through a cross-sectional survey conducted across multiple centers, utilizing a large sample size. The study aims to furnish robust data support for research endeavors in diverse health-related domains within China. It is necessary to conduct a nationwide survey on the mental health status and health behaviors of the population in promptly after the end of the COVID-19 pandemic, which will help us to understand the alterations that the pandemic has brought about in the mental health of Chinese population as well as in their health behaviors. The survey will also help experts and scholars produce more appropriate research results and provide more timely guidance for health care after a pandemic to promote Chinese residents’ health. The survey encompasses a comprehensive range of factors, including physical health status, the influence of diseases on the personal life and work, basic family situation, access to medical care, and participation in public service behaviors.

Additionally, the questionnaire assesses residents’ mental health status, perceived stress capacity, and psychological resilience. It also gathers information on residents’ medication usage, activity levels, dietary habits, sleep patterns, and interpersonal interactions. Furthermore, the survey examines self-rated health status, health literacy, family communication, and childhood experiences. The questionnaire design employed in this study exhibits a comprehensive coverage and breadth of the survey, enabling a thorough examination of our residents’ mental health status and health behaviors.

Alterations in the broader sociocultural milieu can induce modifications in the mental well-being of individuals, particularly among adults, after the cessation of the COVID-19 pandemic(Meaklim et al., 2021). Depression and anxiety, as an essential assessment of mental health, were included in this investigation and the study by Zhu et al.(Zhu et al., 2023) suggested a significant change in the prevalence of depression and anxiety among the respondents during the blockade period, so it was necessary to investigate the depression and anxiety status of the respondents after the full unblocking in this study. In addition, this study also focuses on the psychological resilience of the individual. The results of this study suggest that having good psychological resilience will reduce adverse effects on psychological well-being when negative external events occur(Blanc et al., 2021). Based on the above, in the individual dimension, this study also included the measure of individual’s perception of stress, the measure of the individual’s social support, the measure of an individual’s self-efficacy, the individual’s perception of body image, and the individual’s tendency to take external risks. As for the social dimension, this study measured public attitudes towards people with mental disorders and citizens’ motivation to participate in public services. This study aims to comprehensively explore the mental health status of the Chinese population more comprehensively by measuring of more mental status variables.

The correlation between individual health-related literacy and the preservation of physical and mental well-being becomes notably pronounced during significant social occurrences(Cudjoe et al., 2020). The prevalence of electronic devices and the widespread adoption of electronic screen usage patterns among individuals have underscored the significance of e-health literacy as a crucial determinant of personal health. Research has consistently demonstrated a positive association between e-health literacy and health-related behaviors, with e-health literacy as a potential intermediary in the pathway from health-related information to changes in health-related behaviors(Kim et al., 2023). Adverse childhood experiences significantly impact the emergence of mental health issues in adulthood, with the deleterious consequences of such experiences being transmitted intergenerationally within the family unit via maternal channels (Kang et al., 2021). A high level of drug literacy is fundamental during a pandemic when many residents increase their self-medication behaviors due to lockdowns or the inaccessibility of health services. In a survey of hypertensive older adults, medication-related health literacy was shown to affect long-term safe adherence to medication for people with chronic conditions(Shi et al., 2019).

An individual’s health behaviors, both within their family and social environment, exert significant influence on their physical and mental well-being. Despite focusing on conventional health behaviors such as smoking, alcohol consumption and diet, this study also looks at health-related behaviors that are gaining attention among scholars, such as individuals’ participation in unpaid household chores, public service and participation in the arts. According to the research conducted by Jennifer Ervin et al(Ervin et al., 2023), there is evidence to suggest that prolonged engagement in unpaid domestic work within the household harms individuals’ mental well-being. Furthermore, the allocation of unpaid domestic responsibilities between genders within the household also subtly influences the mental health of both partners involved. A study conducted amidst the COVID-19 pandemic indicates that the engagement of medical patients in social volunteering has the potential to foster a more favorable psychological state(Phillips et al., 2022). In a similar vein, consistent involvement in social volunteering has been found to have a favorable influence on cognitive abilities. In contrast substantial social engagement acts as a protective factor against amygdala deterioration, thereby safeguarding emotionally significant memories and mitigating the likelihood of developing dementia (Carlson, 2020). Studies conducted in the United States and Colombia suggest that participation in the arts can lead to a reduction in the prevalence of depression and anxiety and an increase in the health and well-being of individuals(Bone et al., 2021; Gómez-Restrepo et al., 2022).

This study employed a meticulous questionnaire design and conducted an on-site survey to establish the inaugural survey database on Chinese residents’ mental health and health behaviors in the aftermath of the COVID-19 pandemic. The findings from this database will provide valuable guidance to policymakers and healthcare institutions in formulating more targeted policy recommendations to enhance the physical and mental well-being of Chinese residents post-pandemic. In 2022, a cross-sectional study was conducted to investigate the psychology and behavior factors of Chinese residents, resulting in significant research findings(Li et al., 2023). It is anticipated that the forthcoming study in 2023 will further contribute to the well-being of the Chinese population and global health, as well as advance research in the health field. The release of research protocols improves the transparency of research. It informs the scientific community of what research is being done, which helps to avoid duplication and better coordinate research efforts.

## Author contributions

Writing – Original Draft Preparation, D.L&S.F&X.H&W.G; Writing – Review & Editing, D.L&S.F; Data Curation Y.Y; Supervision, C.L&X.Z&L.S&H.L&F.J&X.Z; Study Deisgn & Supervision & Funding Acquisition & Resources, Y.W&X.S; Project Administration, Others.

## Availability of data and materials

The PBICR project has been ongoing from 2020 to the present, with an annual multi-centre extensive sample size transect survey. We usually distinguish between different years of surveys by the suffix. (e.g., PBICR2023 denotes surveys conducted by PBICR in 2023.)The historical materials and any associated protocols supporting this study’s findings are available from the corresponding authors upon request.

## Declarations

### Research ethics and patient consent

This study (SWYX:NO.2023-198) has been approved by the Ethics Research Committee of Shandong Provincial Hospital. The cover page of the questionnaire will explicitly outline the study’s objectives and ensure the preservation of anonymity, confidentiality, and the participants’ right to decline participation. Informed consent has been duly acquired from all individuals involved in the study. This study was filed in the National Health Security Information Platform (Record No.: MR-37-23-017876) and officially registered in the China Clinical Trials Registry (Registration No.: ChiCTR2300072573)

## Data Availability

All data produced in the present study are available from the corresponding authors upon request.

https://www.x-mol.com/groups/pbicr/shujukukaifang

## Acknowledgments

Not applicable.

## Consent for publication

Not applicable. Availability of data and material.

## Abbreviation

COVID-19: Corona Virus Disease 2019
PBICR: Psychology and Behavior Investigation of Chinese Residents
WHO: World Health Organization

