## Supplementary material for "Study protocol: A national cross-sectional study on psychology and behavior investigation of Chinese residents in 2023, PBICR": Table 1

**Table 1 Scale used for the questionnaire**

| Level | Scale | Abbreviation | Applicable Age | Target Population | Dimension | Item | Score |
|---|---|---|---|---|---|---|---|
| Psychological Level Scale | Big Five Inventory-10 Items[36] | BFI-10 | 12 + years | All Populations | 5 | 10 | 1~5（strongly disagree to strongly agree ). reverse score: questions 1.3.4.5.7, forward score: questions 2.6.8.9.10 Higher scores indicate higher levels of a given personality trait. |
| Psychological Level Scale | Patient Health Questionnaire-9 items[37] | PHQ-9 | 12 + years | All Populations | 1 | 9 | 0~3 (never to nearly every day) 0-4 no depression, 5-9 mild depression, 10-14 moderate depression, 15-19 moderate to severe depression, 20-27 severe depression |
| Psychological Level Scale | Generalized Anxiety Disorder-7 items[38] | GAD-7 | 12 + years | All Populations | 1 | 7 | 0~3 (never to nearly every day) 0 to 4 no anxiety, 5 to 9 mild anxiety, 10 to 13 moderate anxiety, 14 to 18 moderate anxiety, 19 to 21 severe anxiety |
| Psychological Level Scale | Perceived Stress Scale-4 Items[39,40] | PSS-4 | 12 + years | Junior high school or above | 2 | 4 | 0~4 (never to always) with a total score ranging from 0 to 16 points, higher scores indicating more perceived stress. |
| Psychological Level Scale | Perceived Social Support Scale[41,42] | PSSS-SF | 18 + years | All Populations | 1 | 3 | 1~7 (strongly disagree to strongly agree), higher scores indicating greater perception of social support |
| Psychological Level Scale | New General Self-efficacy Scale[41,42] | NGSES-SF | 12 + years | All Populations | 1 | 3 | 1~5 (strongly disagree to strongly agree), with a total score ranging from 3 to 15 points, with higher scores indicating greater self-efficacy |
| Psychological Level Scale | Body Image–Acceptance and Action Questionnaire–5[43] | BI-AAQ-5 | 18 + years | All Populations | 1 | 5 | 1~7(never ture to always ture), the total score range is 5-35 points, higher scores indicate higher levels of body image flexibility. |
| Psychological Level Scale | Connor-Davidson Resilience Scale-2[44] | CD-RISC2 | 18 + years | All Populations | 1 | 2 | 0~4 ( not true at all to true nearly all of the time), with a total score ranging from 0 to 8 points, with higher scores indicating greater resilience. |
| Psychological Level Scale | General Risk Propensity Scale[45] | GRiPS | 18 + years | All Populations | 1 | 8 | 1~5 (strongly disagree to strongly agree), with a total score ranging from 5 to 40 points, with higher scores indicating higher propensity to take risks. |
| Psychological Level Scale | The Stigma-9 Questionnaire[46] | STIG-9 | 18 + years | All Populations | 1 | 9 | 0~3 (disagree to agree), with a total score ranging from 0 to 27 points, with higher scores indicating more stigmatization. |
| Psychological Level Scale | Public Service Motivation Scale-8 items | PSM-SF | 18 + years | All Populations | 4 | 8 | 1~5 (strongly disagree to strongly agree), with a total score ranging from 5 to 40 points, with higher scores indicating higher public service motivation. |

| | | | | | | | |
|---|---|---|---|---|---|---|---|
| Behavior Level Scale | Smoking and Alcohol Behavior Scale | | 18 + years | People over 18 years old | self-developed scale | 11 | |
| Behavior Level Scale | Fagerstrom Test of Nicotine Dependence[47] | FTND | 18 + years | People over 18 years old | self-developed scale | 6 | The total scores ranging from 0 to 10, with higher scores indicating higher nicotine dependence. |
| Behavior Level Scale | The International Physical Activity Questionnaire[48,49] | IPAQ-7 | 18 + years | All Populations | 4 | 7 | ①Walking MET = 3.3 x average daily walking time x weekly walking days.② Moderately strong MET = 4.0 x average time engaged in moderate intensity activity per day x days engaged in moderate intensity activity per week.③Strenuous activity MET = 8.0 x average time engaged in strenuous activity per day x days engaged in strenuous activity per week. Therefore, basal metabolic time per week (minutes) = ① + ② + ③. |
| Behavior Level Scale | Pittsburgh Sleep Quality Index[50] | B-PSQI | 18 + years | All Populations | 5 | 6 | Total score ranges from 0 to 15, with higher scores indicating poorer sleep quality |
| Behavior Level Scale | Community Citizen Behavior Scale-5 items | CCB-5 | 18 + years | All Populations | 1 | 5 | 1~5 (strongly disagree to strongly agree), with a total score ranging from 5 to 25 points, with higher scores indicating more community citizenship behavior. |
| Behavior Level Scale | Intimate Relationship Violence Scale | | 12 + years | All Populations | self-developed scale | 6 | 1~5 (never to almost always), the total score range is 6-30 points, higher scores indicate more intense intimate violence. |
| Behavior Level Scale | Media Use Behavior Scale[52,53] | | 12 + years | All Populations | self-developed questionnaire | 6 | 1~5 (never to always), with a total score ranging from 6 to 30 points, with higher scores indicating more frequent use of media |
| Behavior Level Scale | Antibiotic Use Behavior and Cognition Questionnair | | 12 + years | All Populations | self-developed scale | 17 | 1~5 (strongly agree to strongly disagree ), with a total score ranging from 17 to 85 points. |
| Other scale | Adverse Childhood Experiences[8] | ACEs | 18 + years | All Populations | 7 | 17 | "Yes" or "No", with a total score ranging from 0 to 7 points |
| Other scale | Family Communication Scale[54–56] | FCS-4 | 11 + years | All Populations | 1 | 4 | 1~5 (strongly disagree to strongly agree ), with a total score ranging from 4 to 20 points, with higher scores indicating higher levels of family communication |
| Other scale | Simplified Nutritional Appetite Questionnaire[57,58] | SNAQ | 18 + years | All Populations | 2 | 4 | 1~5 (from "a" to "e"), with total scores ranging from 4 to 20 points, a total score less than 14 indicates that there is a risk of weight loss of more than 5% or 10% in the next months. |

| | | | | | | | |
|---|---|---|---|---|---|---|---|
| Other scale | Morningness-Eveningness Questionnaire-5 items[51] | MEQ-5 | 15 + years | All Populations | 1 | 5 | 1~5, with total scores ranging from 4 to 25. 4 to 7 Definitely Evening, 8 to 11 Moderately Evening, 12 to 17 Neutral, 18 to 21 Moderately Morning, 22 to 25 Definitely Morning. |
| Other scale | Quality of Life Scale[59] | EQ-5D-5L | 18 + years | All Populations | 5 | 5 + EQ-VAS | 1~ 5, with total scores ranging from 5 to 25, with higher scores indicating higher quality of life. Gliding multiple choice questions with EQ-VAS score of 0-100. |
| Other scale | the electronic Health Literacy Scale[60,61] | eHEALS-SF | 18 + years | All Populations | 1 | 5 | 1~5 (strongly disagree to strongly agree), with a total score ranging from 5 to 25 points, with higher scores indicating higher level of electronic health literacy. |
| Other scale | Family Health Scale-Short Form[62,63] | FHS-SF | 18 + years | All Populations | 4 | 10 | 1~5 (strongly disagree to strongly agree), with total scores ranging from 10 to 50 points, with higher scores indicating higher levels of family health |
| Other scale | Health Literacy Scale-Short Form-4[64] | HLS-SF4 | 12 + years | All Populations | 3 | 4 | 0~3 (very difficult to very easy ), with total scores ranging from 0 to 12, with higher scores indicating higher levels of health literacy |
| Other scale | Doctor-patient Communication Preferences Questionnaire | | 12 + years | All Populations | self-developed scale | 7 | |
| Other scale | Medication Literacy Scale | | 12 + years | All Populations | 3 | 6 | 1~5 (always ture to never ture), with total scores ranging from 6 to 30 points. |
